# Incubation periods impact the spatial predictability of outbreaks: analysis of cholera and Ebola outbreaks in Sierra Leone

**DOI:** 10.1101/19003525

**Authors:** Rebecca Kahn, Corey M. Peak, Juan Fernández-Gracia, Alexandra Hill, Amara Jambai, Louisa Ganda, Marcia C. Castro, Caroline O. Buckee

**Affiliations:** Center for Communicable Disease Dynamics, Department of Epidemiology, Harvard T.H. Chan School of Public Health, Boston, MA 02115; World Health Organization, Geneva, Switzerland; Ministry of Health and Sanitation, Freetown, Sierra Leone; World Health Organization Country Office, Freetown, Sierra Leone; Department of Global Health and Population, Harvard T.H. Chan School of Public Health, Boston, MA 02115

**Keywords:** cholera, Ebola, epidemics, modeling, predictability, Sierra Leone

## Abstract

Forecasting the spatiotemporal spread of infectious diseases during an outbreak is an important component of epidemic response. However, it remains challenging both methodologically and with respect to data requirements as disease spread is influenced by numerous factors, including the pathogen’s underlying transmission parameters and epidemiological dynamics, social networks and population connectivity, and environmental conditions. Here, using data from Sierra Leone we analyze the spatiotemporal dynamics of recent cholera and Ebola outbreaks and compare and contrast the spread of these two pathogens in the same population. We develop a simulation model of the spatial spread of an epidemic in order to examine the impact of a pathogen’s incubation period on the dynamics of spread and the predictability of outbreaks. We find that differences in the incubation period alone can determine the limits of predictability for diseases with different natural history, both empirically and in our simulations. Our results show that diseases with longer incubation periods, such as Ebola, where infected individuals can travel further before becoming infectious, result in more long-distance sparking events and less predictable disease trajectories, as compared to the more predictable wave-like spread of diseases with shorter incubation periods, such as cholera.

**Significance statement:** Understanding how infectious diseases spread is critical for preventing and containing outbreaks. While advances have been made in forecasting epidemics, much is still unknown. Here we show that the incubation period – the time between exposure to a pathogen and onset of symptoms – is an important factor in predicting spatiotemporal spread of disease and provides one explanation for the different trajectories of the recent Ebola and cholera outbreaks in Sierra Leone. We find that outbreaks of pathogens with longer incubation periods, such as Ebola, tend to have less predictable spread, whereas pathogens with shorter incubation periods, such as cholera, spread in a more predictable, wavelike pattern. These findings have implications for the scale and timing of reactive interventions, such as vaccination campaigns.

## Introduction

Epidemics of emerging infectious diseases such as Ebola and Zika underscore the need to improve global capacity for surveillance and response (1–3). Forecasting the spatiotemporal spread of infectious diseases during an outbreak can enable responders to stay ahead of an epidemic, but it remains challenging both methodologically and with respect to data requirements. Disease spread is influenced by factors, including: the pathogen’s underlying transmission parameters and epidemiological dynamics; social networks and population connectivity; and environmental conditions (4–7). Previous forecasting efforts have had varying levels of success in predicting the total number of cases and spatiotemporal spread of outbreaks like Ebola, and few have actually been used in real time in the midst of an epidemic (5). Efforts to understand the likely performance of forecasts have shown that heterogeneity in contact structure and number of secondary infections can pose challenges, but reasonable predictions can be made in some cases, depending on disease-specific parameters (4). However, the epidemiological attributes that determine predictability remain poorly defined in real-world settings.

The time taken for individuals to become infectious (the latent period) and symptomatic (the incubation period) following infection, and the relationship between the two, have been shown to play a large role in the epidemic potential of diseases (7–9). In particular, transmission that occurs during the incubation period before an individual develops symptoms can contribute to rapid disease spread. When the latent period is shorter than the incubation period for an infectious individual, pre-symptomatic transmission can be a strong driver of the total number of secondary infections by an infectious individual in a completely susceptible population (i.e. R_0_) (8, 9). Indeed, the basis of contact tracing protocols during an outbreak reflect the need to identify and contain individuals during the incubation period, and the relative effectiveness of interventions such as symptom monitoring or quarantine significantly depends on the relationship between infectiousness and symptoms (9). The incubation period is also likely to play a particularly important role in determining the spatial spread of an epidemic because one’s typical travel may continue prior to symptom onset, whereas travel behavior may change or stop altogether during illness (10), particularly when symptoms are severe or immobilizing.

Back-to-back epidemics of cholera (2012-2013) and Ebola (2014-2015) in Sierra Leone present a unique opportunity to compare the spatial dynamics of two epidemics in the same population caused by pathogens with notable similarities in both the drivers of outbreaks and the interventions used to curtail them, including oral rehydration (11, 12). Both are transmitted through contact with contaminated diarrhea or vomitus (plus other bodily fluids for Ebola), and the reproductive number (R_0_) for both diseases is thought to be between 1 and 3 (13, 14). Both diseases can cause immobilizing gastrointestinal symptoms of diarrhea and vomiting and, untreated, their case fatality rates can exceed 50% (15, 16). Cultural factors and rituals, such as traditional funeral practices, are known to influence the spread of both cholera (17) and Ebola (18), while water, sanitation, and hygiene (WASH) programs are often used to slow the spread of each (19). Both epidemics occurred against a backdrop of an immunologically naïve population. Presumably, travel patterns and the density and distribution of people were broadly similar over the time period in question. One critical difference between the dynamics of these diseases, however, is the incubation period, which is estimated at a median of 8-12 days between infection and onset of symptoms for Ebola (1) and only 1-2 days for cholera (20).

We hypothesize that the disease incubation period may be a particularly influential driver of different patterns of disease spread through space and time. We analyze the spatiotemporal dynamics of a cholera outbreak and an Ebola outbreak in Sierra Leone, both of which occurred over a similar time period. We develop a simulation model of the spatial spread of an epidemic and examine the impact of the incubation period on the dynamics of spread and the predictability of outbreaks. We find that differences in the incubation period alone can determine the limits of predictability for these diseases with different natural history, both empirically and in our simulations. Our results show that diseases with longer incubation periods, such as Ebola, where infected individuals can travel further before becoming infectious, result in more long-distance sparking events and less predictable disease trajectories, as compared to the more predictable wave-like spread of diseases with shorter incubation periods, such as cholera.

## Results

We first summarize the cholera and Ebola epidemics in terms of their dynamics in time and space. More cases were reported during the cholera epidemic (22,691) than during the Ebola epidemic (11,903); however, far fewer cholera cases were fatal (324 vs. 3,956). Both epidemics lasted for similar periods of time, with cholera (January 7, 2012 – May 14, 2013) occurring two years prior to Ebola (May 18, 2014 – September 12, 2015). The times between the onset of an outbreak and when half or all of its cases were reported were longer when outbreaks were aggregated by district instead of chiefdom (Figure 1), which has implications for the optimal scale for surveillance and response measures. The median time for a chiefdom cholera outbreak to report half its case total was 3.9 weeks, and median outbreak duration was 11.3 weeks. The median time for district outbreaks to report half their cholera cases was 7.9 weeks, and the median outbreak duration was 43.7 weeks. Analysis of Ebola revealed similar trends, with chiefdoms reporting half of their cases at a median of 13.9 weeks and median outbreak duration of 43.3 weeks, and districts reporting half of their cases at a median of 23.5 weeks and median outbreak duration of 64.1 weeks.

**Figure 1.**
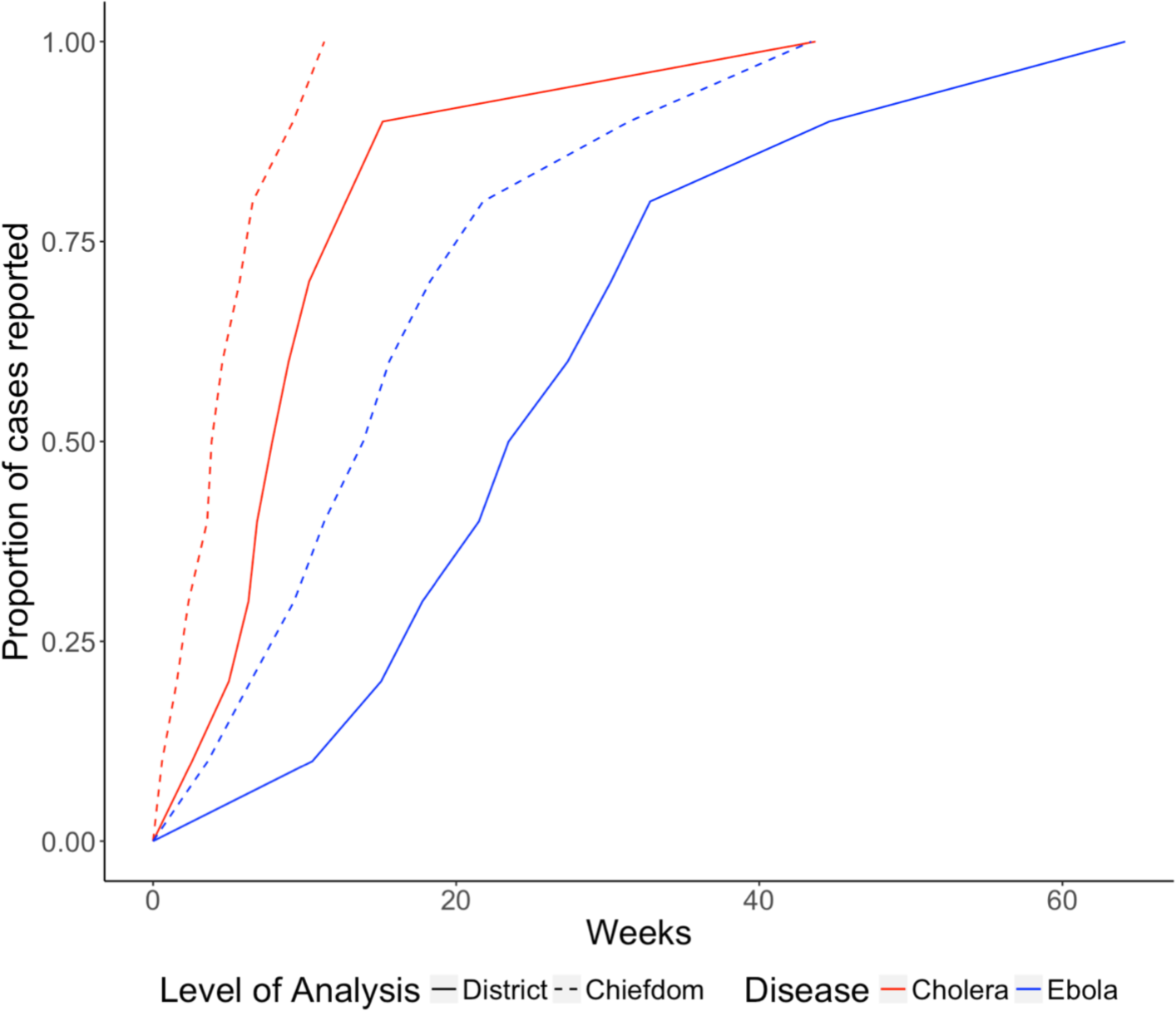
The proportion of cholera and Ebola cases reported over time differed between district and chiefdom level. The times between the onset of an outbreak and when half or all of its cases were reported were longer when outbreaks were aggregated by district instead of chiefdom, which has implications for the optimal scale for surveillance and response measures. The median time for a chiefdom cholera outbreak to report half its case total was 3.9 weeks and a median of 7.9 weeks for district cholera outbreaks. For Ebola, chiefdoms reported half of their cases at a median of 13.9 weeks and districts at a median of 23.5 weeks.

Both the cholera and Ebola epidemics were widespread, each reaching more than 75% of the country’s chiefdoms. However, their trajectories differed. The spread of cholera from the northwest followed a radial spatial dispersion gradually in all directions for the first six months, while Ebola spread from the southeast for two months before rapid expansion to the northwest which sparked the national epidemic (Figure 2 A-B). These findings were statistically supported by space-time analysis of each epidemic, which revealed clusters of high case reporting of both diseases in western Sierra Leone and unique clusters of cholera in the south and Ebola in the east (Figure S1). The wave front of chiefdom cholera outbreak onset progressed more slowly and gradually than for Ebola, which exhibited faster and more discontinuous expansion as shown by the larger spacing between monthly contour lines (Figure 2 A-B). Despite their different trajectories, the geography of the epidemics largely overlapped, with clusters of high cumulative attack rates of cholera and Ebola observed in the north and west regions of Sierra Leone (Figures 2 C-D) and confirmed through Local Moran’s I methods (Figure S2).

**Figure 2.**
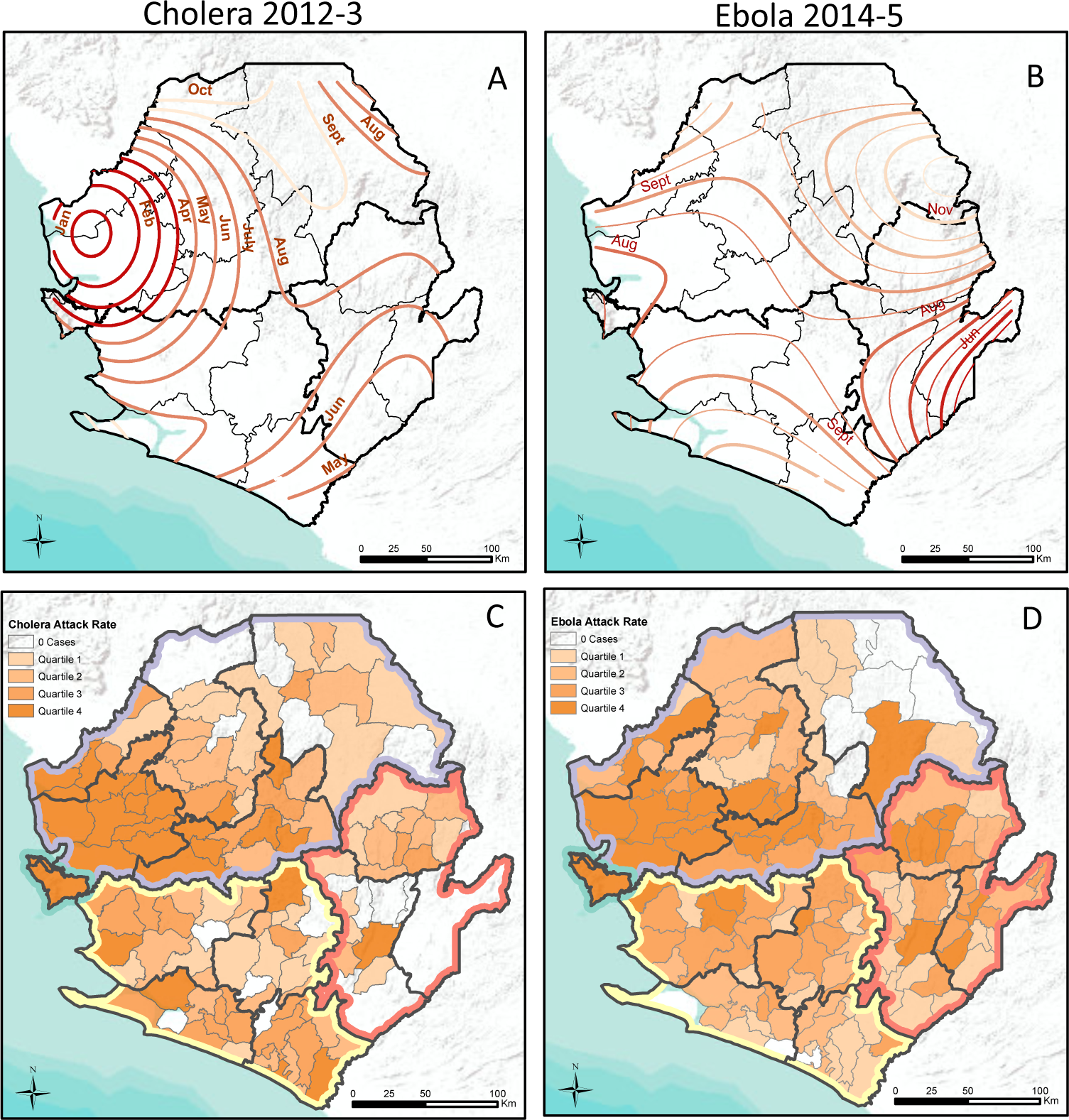
Spatial trend contours of disease spread and chiefdom attack rates highlight similarities and differences between the two epidemics. Spatial trend contours of cholera (A) and Ebola (B) spread from areas in dark red to light red; thicker lines (A & B) show monthly increments and thinner lines (B) show 2 week increments. Thick black lines denote regions and thin black lines denote districts. Chiefdom attack rate quartile for cholera (C) and Ebola (D) vary over space and regions. Colored boundaries denote regions, followed by bold black borders for districts and thin borders for chiefdoms.

As a daily estimate of transmission intensity, we recorded the effective reproductive number (R_t_) and its variation over time nationally and by region (Figure 3). While some areas sustained transmission (i.e., R_t_ > 1) of both cholera and Ebola for many days (e.g., Freetown in the west and Kenema Town in the east), most chiefdoms recorded either zero cases or zero days with R_t_ > 1 (Figure S3). As expected, transmission intensity of both diseases was positively correlated in chiefdoms near each other (Figure S4). Correlation decayed with distance, consistent with local disease spread, and inter-chiefdom distances of over 100km eliminated any evidence of positive correlation of disease presence, chiefdom outbreak time, case count, and cumulative attack rate (Figure S4). These metrics appear more highly correlated in space for cholera than for Ebola, although the confidence intervals overlap.

**Figure 3.**
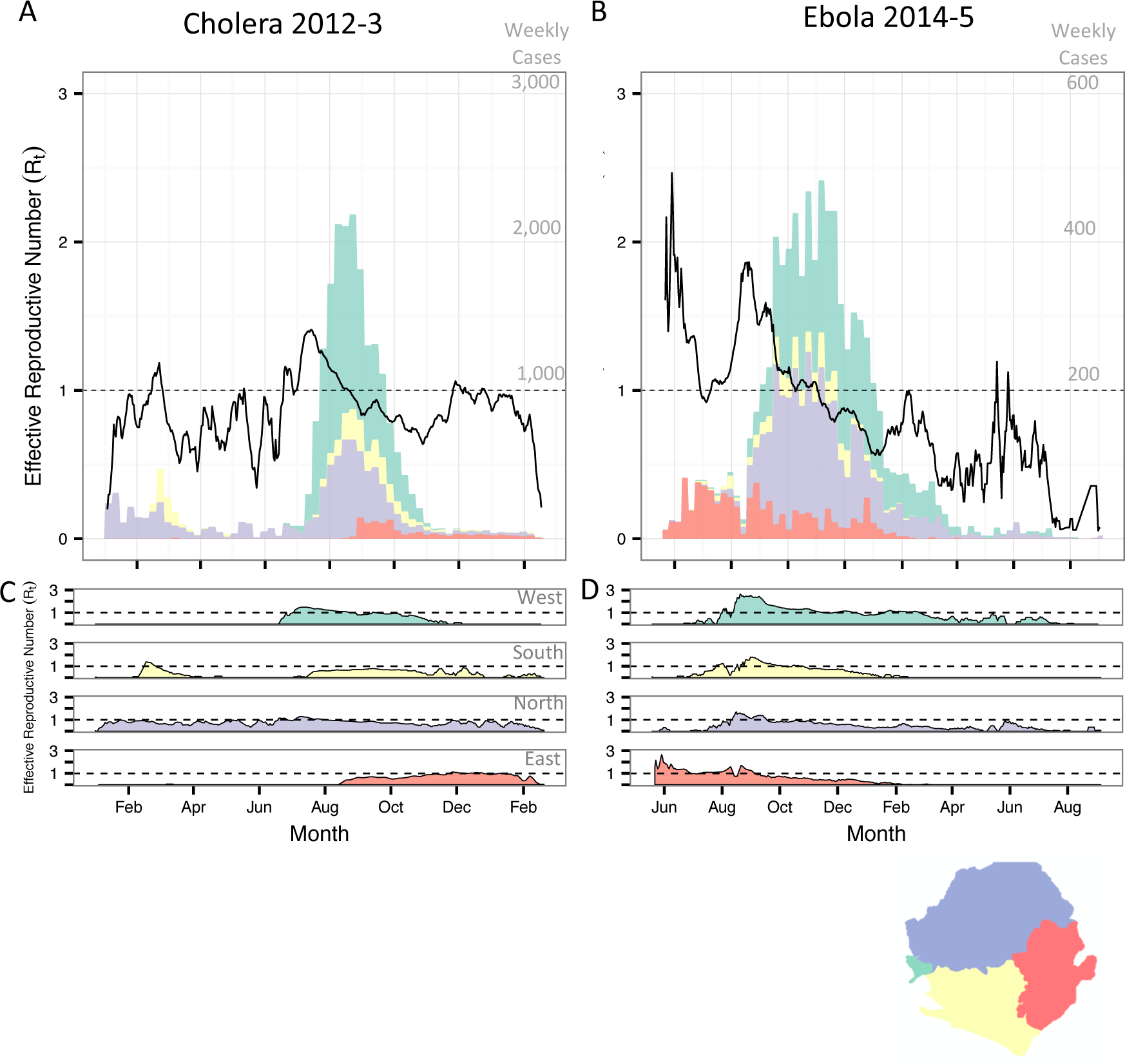
Weekly case counts show outbreak trajectory in the four regions of the country. The bars in A and B indicate the weekly case count on independent y-axes of cholera and Ebola, respectively. Black lines show maximum likelihood estimates of R_t_ of cholera and Ebola epidemics nationally (A and B, respectively) and in each region (C and D, respectively).

### Simulations

Our simulations show a systematic relationship between the incubation period and spatiotemporal patterns of disease spread. As expected, simulated epidemic curves of diseases with shorter incubation periods were more acute while diseases with longer incubation periods peaked later (Figure 4A). Although epidemics tend to last longer for diseases with longer incubation periods, the spread of the disease to more distant locations can progress more quickly, causing a discontinuous and more rapidly spreading wave front (Figure 4 C-D). In the first 50 days of our simulations, locations further from the origin of the epidemic experienced cases earlier on average in simulations with longer incubation periods compared to those with shorter incubation periods, likely due to long-distance sparking events from infected agents traveling during the incubation period (Figure 4B). The dispersion kernel Kx(d), the probability that an agent will end up at a position separated a distance d from the initial position after x days, is more homogeneously spread and has non-vanishing probabilities at greater distances the higher the incubation period, explaining the enhancement in sparking events (Figure S5).

**Figure 4.**
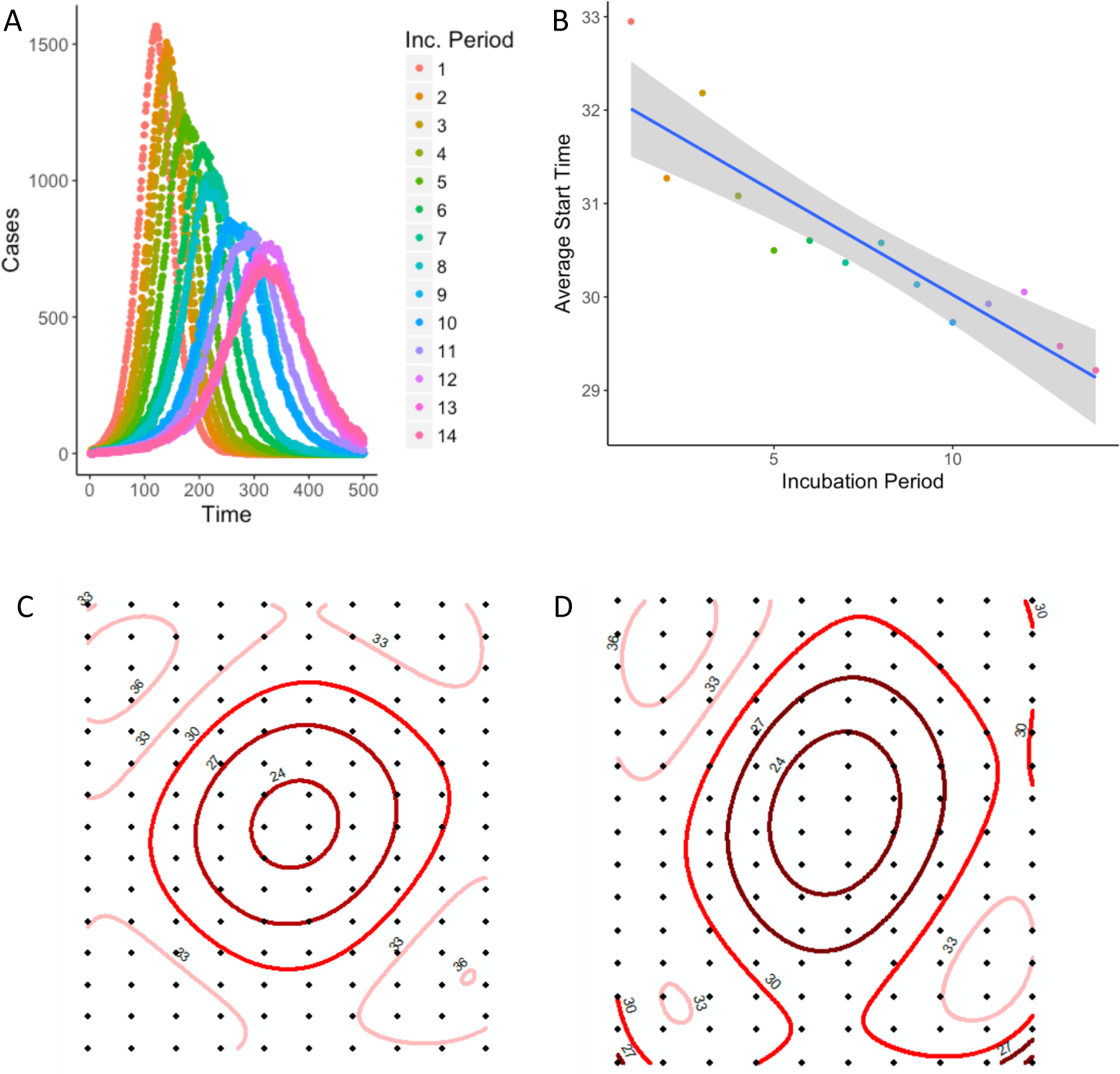
Results of 700 simulations of 14 different incubation periods show the impact of incubation period on disease spread. Epidemics with shorter incubation periods are more acute than epidemics with longer incubation periods (A). The average start time of epidemics at all locations over the first 50 days of outbreak is later for shorter incubation periods than longer (B). Spatial trend contours of first 50 days of simulated outbreaks with shorter incubation period (2 days) (C) and longer incubation period (10 days) (D), spreading from areas in dark red to light red, show that shorter incubation periods result in a more wave front spread and longer incubation periods result in more long-distance sparking events; numbers show average start day relative to start of the outbreak.

Simulations on a lattice with relative population size based on Sierra Leone’s chiefdom census data support the finding that the duration of epidemics is longer on a district (i.e. group of lattice points) rather than chiefdom (i.e. individual lattice point) scale, with duration lengthening with increasing incubation periods (Figure 5A).

**Figure 5.**
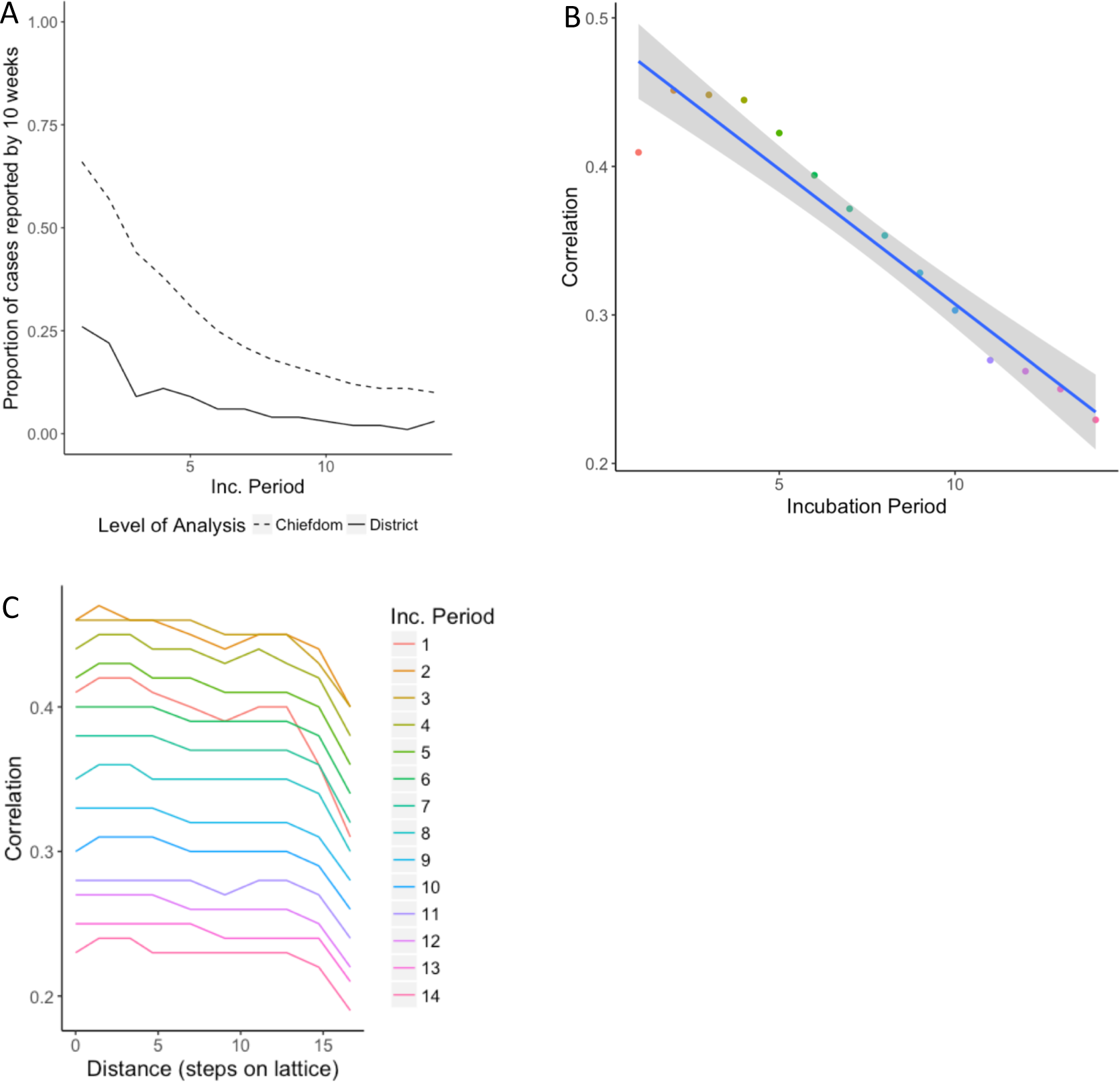
The incubation period impacts the timing of outbreaks and as a result, the correlation. As the incubation period increases, the proportion of cases reported by 10 weeks, when a reactive vaccination campaign might begin, decreases in simulated epidemics (A). As the incubation period increases, the average correlation overall (B) and by distance from origin of simulated outbreaks (C) decreases.

Consistent with the correlation analysis comparing Sierra Leone’s cholera and Ebola outbreaks, time series from simulated outbreaks with shorter incubation periods were more highly correlated than those from simulations with longer incubation periods, with correlation decaying as distance between locations on the lattice increased (Figure 5 B-C). Higher correlation suggests increased predictability, which the results of the overlap function support (Figure 6). As the incubation period lengthened, the average predictability during the beginning of the outbreak decreased as the epidemics spread via unpredictable sparking patterns. Predictability plateaued as the outbreaks became widespread.

**Figure 6.**
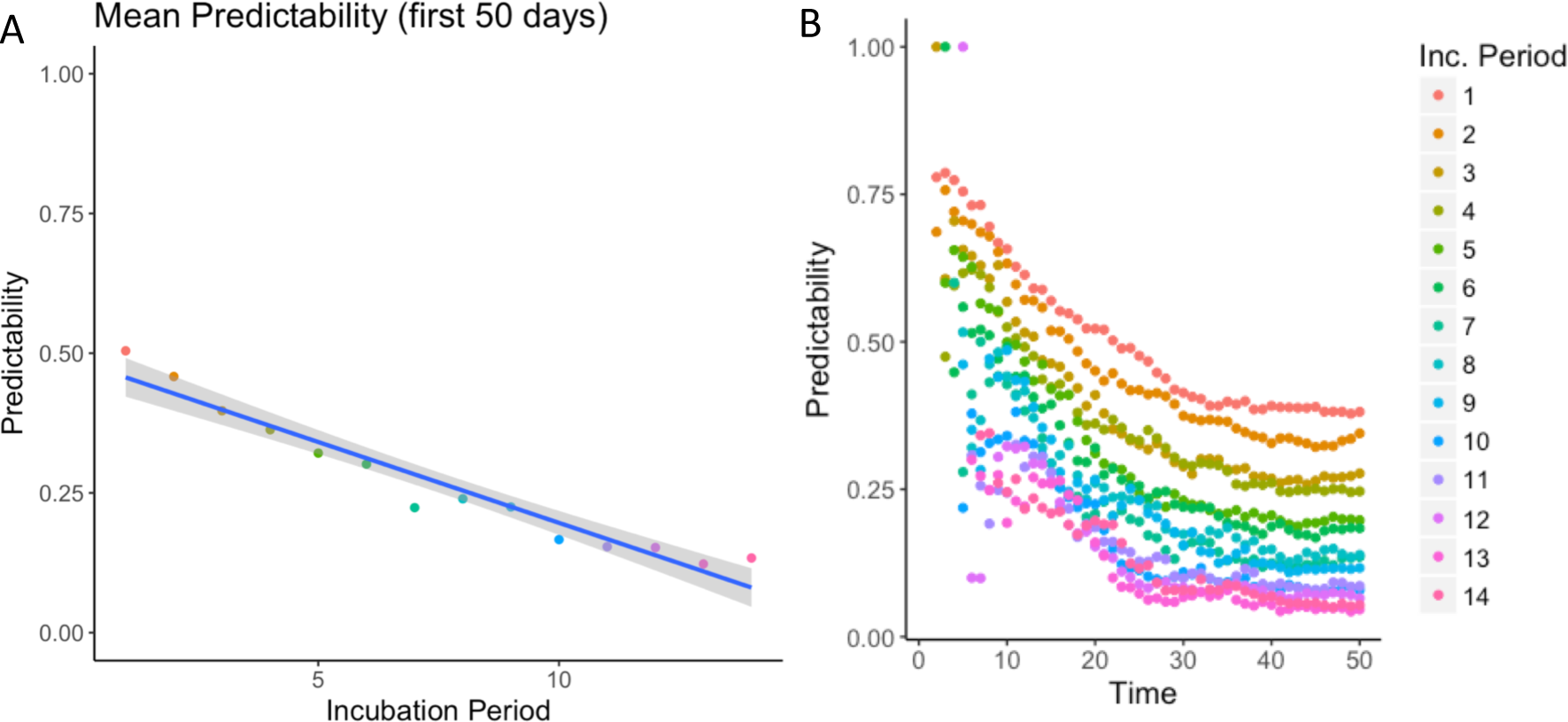
The incubation period impacts the predictability of disease spread. As the incubation period increases, the average overlap (predictability) of the first 50 days (A) and over the first 50 days (B) of simulated outbreaks decreases.

## Discussion

Analysis of the cholera and Ebola epidemics revealed commonalities and differences in the way these pathogens spread throughout Sierra Leone, and our simulations suggest the differences in the incubation period reproduce these differences. Spatial diffusion of Ebola occurred more quickly than cholera, as evidenced by the wave front contour lines and further supported by statistical tests considering a subset excluding cholera cases before the brief respite in June (Figure S6). Additionally, cholera metrics were more correlated in space than Ebola metrics. Our model simulations suggest that these findings are potentially due to the counter-intuitive role of the longer incubation period for Ebola as compared to cholera. Travel during the incubation period will be a key driver of geographic disease dispersion and predictability, especially in a population of individuals who decrease mobility when ill. Consequently, diseases with longer incubation periods will tend to have more long-distance sparking events caused by infected, but healthy, individuals traveling during the incubation period. This will result in faster epidemic dispersion to distant, unpredictable locations. These findings are in line with Marvel et al.’s results, which found epidemic wave fronts are less likely to occur for mobility kernels that decay more slowly (21); when the incubation period is longer, the effective kernel can span to more distant places, making sparking events more probable.

Similar results were also obtained when infectious agents did not decrease mobility when ill, suggesting that travel during the incubation period has more influence on correlation and predictability than travel during the infectious period. While many other factors will influence wave speed, continuity, and epidemic synchrony, our simulations showed that small changes in the incubation period can powerfully influence epidemic dynamics.

The incubation period has already been recognized as an important component for understanding epidemics and control (8), with the conventional knowledge that long incubation periods allow more time for responders to scale-up interventions against the overall epidemic and are therefore advantageous for disease control efforts. Here we demonstrated a counter- intuitive mechanism whereby a longer incubation period may in fact hinder a response by decreasing the predictability of outbreaks and increasing their geographic scope as well as of the needs of surveillance and response. We use simulations to reproduce the double-edged sword of the influence of the disease incubation period on reactive interventions.

Reactive vaccination strategies exist for both cholera and Ebola outbreaks, and a better understanding of spatiotemporal spread can facilitate locally-preemptive vaccination to target locations at high risk of introduction (22–24). Reactive vaccination campaigns must consider both the expected duration of an outbreak at a given spatial scale and the predictability of its spread. For cholera, we showed that chiefdom outbreaks tended to report half their cases within approximately 4 weeks, suggesting reactive vaccination of a chiefdom triggered by detection of a case may not be early enough to avert an outbreak and instead intervening at a wider scale, such as districts, might provide more favorable timing for intervention targeting. We posit for future study that regional-ring vaccination strategies may be better suited to diseases with short incubation periods, while contact-ring vaccination strategies may be better suited to diseases with longer incubation periods due to their regional unpredictability and the longer intervals between generations in infection.

There are limitations to our work with regards to data as well as methods. Few cholera cases were confirmed during the epidemic and therefore we depend on the clinical definition as well as the cases that were detected and recorded by the surveillance system. Ebola surveillance data is similarly prone to differences in reporting rates, but the use of only confirmed cases yielded similar results. Our estimates for the effective reproductive number depend on, and absorb the limitations of, case data, serial interval estimates, and the chiefdom connectivity matrix. Specifically, we assume all cases in our dataset acquired infection from others in the dataset, thereby excluding missing cases and asymptomatic transmitters. However, this method has been shown to be robust to cases missing at random and we furthermore expect the role of asymptomatic transmission to be limited for both diseases due to the strong correlation between pathogen load, symptoms, and infectiousness (25, 26).

Further, we assume no changes to the serial interval for either cholera or Ebola during the course of the epidemics. For cholera specifically, waterborne transmission could potentially lead to a heavy right-tail in serial intervals or change the distribution as pathogen accumulates or clears from a drinking source. Household data in Bangladesh, where the role of water contamination is expected to be large, suggest few serial intervals beyond 7 days (27). The geographic spread of cholera in Sierra Leone from the northwest and south towards the center of the country was not consistent with the direction of key waterways in the country, which primarily run from the eastern highlands to the western shores, suggesting population density and human-to-human contact likely played a larger role than water sources in this outbreak.

Finally, our simulation model provides a proof-of-concept test of the hypothesis of the impact of the incubation period on disease spread and makes several simplifying assumptions. These assumptions could be relaxed in future work, including the complete overlap of symptoms and infectiousness and constant diffusion of agents without increased probability of returning “home.” Other models for connectivity and movement could also be explored.

The threat of cholera and Ebola re-emergence in Sierra Leone remains a concern (28). We have shown that differences in incubation period alone are a powerful driver of geographic dispersion and merit further study. Although this study only examines one epidemic from each disease, the size of these epidemics, combined with simulation results from our model, can lend information towards a better understanding of each disease and our ability to predict disease spread. This work can inform development of international preparedness and response strategies and ensure timely and effective interventions.

## Methods

### Data

Cholera cases were reported to the Sierra Leone Ministry of Health and Sanitation by treatment facilities throughout Sierra Leone between January 1, 2012 and May 15, 2013. Following standard WHO definitions (29), a suspected cholera case was defined as acute onset of watery diarrhea or severe dehydration in a person aged five years or older in a region without a known cholera outbreak; once the Government of Sierra Leone declared an outbreak of cholera on February 27, 2012, any case of acute watery diarrhea could henceforth be included as a suspected cholera case. Data were compiled and anonymized by the WHO for analysis, with case reports temporally resolved by day and spatially resolved by chiefdom. For Ebola, we used a published dataset of 8,358 confirmed and 3,545 suspected Ebola cases reported to the Sierra Leone Ministry of Health and Sanitation from May 2014 to September 2015 (30). Our analysis included both suspected and confirmed cases of Ebola according to standard WHO definitions (31). Population estimates for 2012 and 2014 were imputed by chiefdom using a linear fit between chiefdom population estimates from the 2004 and 2015 Population and Housing Censuses (32).

Sierra Leone has four administrative regions, which are divided into fourteen districts. Freetown, the capital and largest city, is comprised of two districts; the remaining twelve districts are subdivided into 149 chiefdoms, with a median of 11.5 chiefdoms per district. Chiefdom, as the finest administrative unit available for cases of both cholera and Ebola, was considered the unit of observation and the unit of analysis (with the exception of cases in Freetown which were solely reported at district level), as it is the likely scale of intervention campaigns like vaccination. To understand what would have been observed at a coarser spatial scale that is more common for surveillance, we additionally aggregated cases by district.

### Spatiotemporal analysis

We defined the first outbreak week for each chiefdom as the week of the first reported case in that chiefdom. We visualized outbreak spread using a contour map of outbreak wave front direction and speed (30). Contours of spatial spread were generated using ArcMap 10.3.1 Spatial Analyst extension by applying a fourth degree polynomial trend interpolation of chiefdom onset dates and generating contour lines of this surface in 2–4 week increments. With this method, more closely-spaced contour lines indicate slower propagation, similar to the slope of a topographic map of geographic elevation.

To identify space-time clusters, using the SaTScan software package (33), we ran a retrospective discrete Poisson-based Scan Statistic over the entirety of the outbreaks for which data were available, namely 16 months of cholera data and 17 months of Ebola data. Disease case reports were assumed to be Poisson-distributed given chiefdom population size. The unit of time aggregation for the analysis was specified as the median incubation periods for each disease (1.5 days for cholera and 10 days for Ebola).

We calculated spline correlograms for four chiefdom outbreak metrics to measure spatial correlation of date of first case, case count, attack rate, and disease presence (yes/no). The maximum centroid-to-centroid distance was set to 150 km, approximately the radius of Sierra Leone. We used the spline.correlog function of the R package *ncf* for each disease and all chiefdom pairs (34).

We estimated the daily effective reproductive number (R_t_), the average number of onward infections generated by cases with onset on day *t*, using methods described by Wallinga and Teunis and extended to metapopulations by White *et al*. (35, 36). This maximum likelihood method estimates the probability that an observed case was the infector for each subsequent case by leveraging information on the daily case count, the serial interval distribution (i.e., the time between symptom onset of an infector-infectee pair), and a weights matrix that quantifies relative contact frequency within and between chiefdoms. The serial interval for cholera was assumed to follow a gamma distribution (rate = 0.1, shape = 0.5) with a median of five days, as has been used previously after consideration of both fast, person-to-person, and slow, environmental, transmission routes (37, 38). The serial interval for Ebola was assumed to follow a gamma distribution (rate = 0.17, shape = 2.59) with a median of 13.3 days derived from the estimates by the WHO Ebola Response Team (1). The contact frequency between two given chiefdoms was assumed to decrease with squared distance between the chiefdom centroids. Additional weights matrices with different functional forms for distance decay yielded qualitatively similar measurements of R_t_.

### Model

We simulated an agent-based model with 45,000 agents distributed equally in 150 locations, evenly spaced on a 15 × 10 lattice. Infected agents progressed through a traditional Susceptible-Exposed-Infectious-Recovered (SEIR) compartmental transmission framework. We assumed the incubation period (i.e. the time from exposure to symptom onset) overlapped completely with the latent period (i.e. the time from exposure to onset of infectiousness). Similarly, the duration of illnesses (5 days) aligned with the duration of infectiousness. Movement of agents between two locations was based on a gravity model, whereby connectivity was proportional to the population sizes of each location and the inverse squared distance between them (39). Different parametrizations of the gravity model, as well as simulations with relative population size based on Sierra Leone’s chiefdom census data (40), yielded similar results.

Susceptible, exposed, and recovered individuals had a daily probability of movement. To simulate the impact of a reduction in mobility during illness, agents in the model had their movement reduced as far as zero throughout the course of their period of infectiousness (and, equivalently, illness). Holding all other parameters constant, we conducted 700 simulations of epidemics for incubation periods ranging from 1 to 14 days. We seeded the epidemic at the same location near the center of the lattice for all simulations.

Synchrony was assessed with the R package *ncf* functions mSynch and Correlog.Nc (34), which both estimate the correlation between the time series in each of the 150 locations across the 500 days of the simulations, with the latter incorporating distance (34). which both estimate the correlation between the time series in each of the 150 locations across the 500 days of the simulations, with the latter incorporating distance. To assess the impact of the incubation period on the initial speed of spread, we calculated the average start time across all locations in the first 50 days of the outbreaks as well as at increasing distances from the location on the lattice where the outbreaks began.

To estimate the predictability of outbreak spread in space and time, we adapted an overlap function used to measure predictability of a SARS outbreak (7). In each simulation, a vector *π*_*j*_(*t*) represents the proportion of all infected individuals at time (t) who are at location (j). In a system with high predictability, *π*_*j*_(*t*) will be similar across simulations. The overlap between simulations I and II can be estimated by: 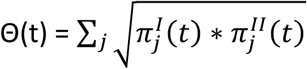. Θ(*t*) ranges from 0 to 1 with a higher value indicating more overlap and thus more predictability. We estimated predictability at each time point by calculating the average of the overlap functions for each pair of simulations for each incubation period. We calculated the average overlap across time points to provide a summary metric for predictability of each incubation period. Code and data are available on Github (41).

## Data Availability

Code and data are available on Github: https://github.com/rek160/Sierra-Leone-Cholera-Ebola

## Acknowledgements

The authors thank Dr. Foday Dafae for early support of this work.

This work was supported by award number U54GM088558 from the National Institute of General Medical Sciences. The content is solely the responsibility of the authors and does not necessarily represent the official views of the National Institutes of Health.

